# A Computational Model for Estimating the Progression of COVID-19 Cases in the US West and East Coasts

**DOI:** 10.1101/2020.03.24.20043026

**Authors:** Yao-Yu Yeo, Yao-Rui Yeo, Wan-Jin Yeo

## Abstract

The ongoing coronavirus disease 2019 (COVID-19) pandemic is of global concern and has recently emerged in the US. In this paper, we construct a stochastic variant of the SEIR model to make a quasi-worst-case scenario prediction of the COVID-19 outbreak in the US West and East Coasts. The model is then fitted to current data and implemented using Runge-Kutta methods. Our computation results predict that the number of new cases would peak around mid-April 2000 and begin to abate by July, and that the number of cases of COVID-19 might be significantly mitigated by having greater numbers of functional testing kits available for screening. The model also showed how small changes in variables can make large differences in outcomes and highlights the importance of healthcare preparedness during pandemics.

**Author Summary:** Coronavirus disease 2019 (COVID-19) has escalated into a global pandemic and has recently emerged in the US. While some countries have managed to contain COVID-19 efficiently, other countries previously thought to have been well-prepared for outbreaks due to higher living standards and healthcare quality have witnessed an unexpected number of cases. It is currently unclear how the US can cope with the COVID-19 pandemic, especially after mishaps during the initial stages. Our study combines conditions unique to the US and transmission dynamics in regions affected most by COVID-19 to produce a quasi-worse-case scenario of COVID-19 in the US and shows the importance of healthcare preparedness during pandemics.

## Introduction

Coronaviruses (CoVs) comprise a family of enveloped positive-stranded RNA viruses that are known to infect a broad range of animals ranging across mammals and birds. CoVs have been prevalent worldwide for several decades [8, 26] and cause diseases generally associated with the respiratory, gastrointestinal, hepatic, and nervous systems. To date, there are seven CoVs that are known to infect humans and are associated with respiratory symptoms. Four CoVs (HCoV-229E, HCoV-NL63, HCoV-OC43, and HKU1) present relatively mild respiratory tract infections [26], whereas the other three CoVs, Severe Acute Respiratory Syndrome-related Coronavirus (SARS-CoV), Middle East Respiratory Syndrome-related Coronavirus (MERS-CoV), and the novel Severe Acute Respiratory Syndrome Coronavirus 2 (SARS-CoV-2) are considerably more virulent and have caused outbreaks with pandemic potential. They are responsible respectively for the 2002-2003 SARS outbreak [6, 15], the 2012 MERS outbreak [36], and the ongoing COVID-19 pandemic [37]. As CoVs have a global distribution, exhibit considerable genetic diversity and genomic recombination, and have zoonotic potential while sympatry between humans and wildlife increase [5, 26], it is very likely that there will be new CoV outbreaks in the future.

The ongoing COVID-19 pandemic (previously 2019-nCoV) originated from Wuhan city in Hubei province of China [37]. While the first case was reported in December 2019, it has since been thought to emerge as early as November 2019 [25]. Since then, COVID-19 has continued to spread around the world, and at this date, over 150 countries have been affected [3]. Although some countries have managed to contain COVID-19 efficiently, others previously thought to have been well-prepared for outbreaks due to higher living standards and healthcare quality have witnessed an unexpected number of cases [24]. As a result, the scale of COVID-19 within each nation has become relatively uncertain and has only heightened social and economic unrest.

COVID-19 was first reported in the US on January 20, 2020 [11]. While little initial action was taken during the initial days, an exponential increase in the number of cases [24] spurred immediate actions in an attempt to contain COVID-19. For instance, social distancing is being enforced via the closure of educational institutions, restrictions on travel, and suspension of events, and research funding for SARS-CoV-2 has increased. Unfortunately, containment of COVID-19 has been hindered by various factors, such as the initial production of defective test kits [27], a current limited availability of test kits [28, 29], and lack of medical supplies [30, 31]. It is currently unclear how well the US healthcare system will cope with the COVID-19 pandemic over the next few months, especially in view of the lack of adequate projections of the scale of COVID-19 infections and mortality across the country.

In this paper, we attempt to construct a mathematical model that simulates the scale of COVID-19 outbreak in the US. Despite the uncertainty of the pathogenicity and molecular virology of SARS-CoV-2 [23], a few key features pertaining to its transmissibility have been studied. The basic reproduction number *R*_0_, which represents the average number of new infections an infectious person can cause in a naïve population, generally range from 2.3 to 2.9 [22, 33, 38]. The incubation period (the time between initial exposure and development of symptoms) is approximately 5.2 days on average [16, 19], and the infectious period (the time from onset of symptoms to isolation) is approximately 3 days on average [20, 21]. These transmission features are useful parameters to consider for modeling, which we will describe in detail next.

## Methods: Designing the Model

To model the COVID-19 outbreak, we use a variant of the SEIR model (see [7, 18] for some examples). We mainly focus on the East and West Coast of the United States. We assume that there is no travel between the different population zones, and that the natural birth and death rates are equal.

Figure 1 depicts the basis of our model, focusing on a particular coast, where arrows indicate the flow of the population at different stages.

**Figure 1:**
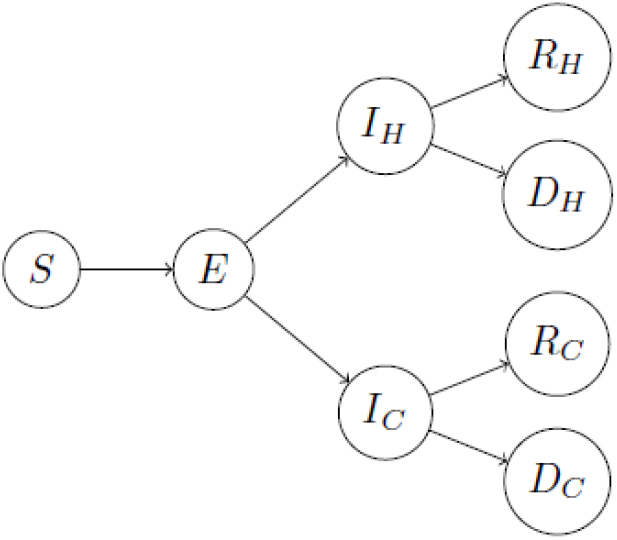
A diagram summarizing our modified SEIR model for each coast.

In Figure 1, *S* represents the susceptible population, and *E* represents the exposed population (i.e. individuals who have been infected but are not yet themselves infectious). The infected population *I* divided into two groups, *I*_*H*_ and *I*_*C*_, wherein the subscripts *H* and *C* stand for “hospital” and “community” respectively. *I*_*H*_ represents those that are infected and isolated (such as those that have been pre-tested and found to carry the virus), and *I*_*C*_ for those that are infected but not isolated (such as those with unreported cases or present mild symptoms that are overlooked). Thus, people in *I*_*H*_ is unable to spread the virus whereas those in *I*_*C*_ can spread the virus. As testing kits in the US are currently in low supply, the current model projects significantly higher levels of *I*_*C*_ than *I*_*H*_ at any given point in time. Once infected, there are two possibilities: either recovery or death. These outcomes are represented by the populations *R* and *D* above, with subscripts as designated above. We also assume the recovered population will have acquired immunity to the virus and are no longer susceptible.

In the mathematical model underlying Figure 1, we treat the various symbols above as functions over time, and the precise variables for ordinary differential equations are as follows.

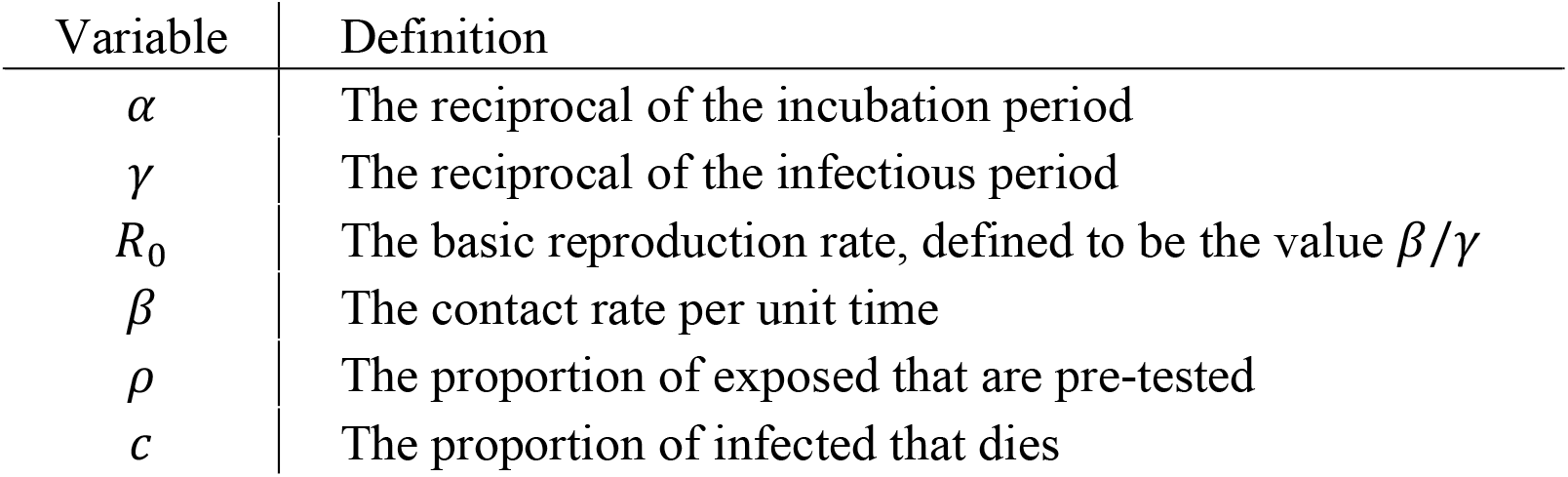

| Variable | Definition |
| --- | --- |
| $\alpha$ | The reciprocal of the incubation period |
| $\gamma$ | The reciprocal of the infectious period |
| $R_0$ | The basic reproduction rate, defined to be the value $\beta/\gamma$ |
| $\beta$ | The contact rate per unit time |
| $\rho$ | The proportion of exposed that are pre-tested |
| $c$ | The proportion of infected that dies |

The expression *N* = *S* + *E* + *I*_*H*_ + *I*_*C*_ + *R*_*H*_ + *R*_*C*_ represents the *effective total population* in the US, i.e. the population that matters for the purposes of virus transmission. Then, for each coast in the US, the discussion above can be mathematically modeled as follows. 

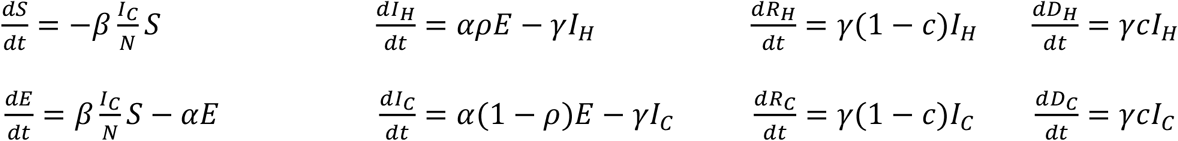

Notice that *R*_0_ is not present in the model above. However, *R*_0_ is important as the contact rate is hard to directly estimate, and we will use *R*_0_ and *γ* to estimate *β* using the relation *β* = *R*_0_*γ*. The basic reproduction rate is also not static over time, and we will use different values over the following three phases:

- Phase 1: There is no action done on the epidemic
- Phase 2: Some action is done to slow down the epidemic, and the coast is preparing for the next phase.
- Phase 3: Coast shutdown; schools moved online, events canceled, and most public areas closed.

To prepare for a quasi-worst-case-scenario, we assume *R*_0_ ≥ 1. This is because *R*_0_ = 1 represents a neutral reproduction rate, whereas *R*_0_ < 1 and *R*_0_ > 1 represents less and more spread, respectively. The next section describes in detail how we simulate our model.

## Methods: Implementing the Model

We simulated our model in MATLAB to prepare for a quasi-worst-case-scenario. We employed a fourth-order Runge-Kutta method [12] to numerically solve the above specified ordinary differential equations, with the range *t* ∈ [0, 250] and stepsize 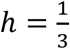 (both measured in days). As for the initial conditions for each coast, we assume that there will not be any pre-existing immune responses that may help defend against the virus, due to SARS-CoV-2 being sufficiently divergent from other CoVs [23]. Hence, the entire population is initially susceptible due to the virus. For example, if a single infected person is introduced at *t*_0_ = 0 to a population of 53 million people (e.g. West Coast US, including Seattle), we will then have *S*(*t*_0_) = 53 * 10^6^, *I*_*c*_(*t*_0_) = 1, and *E*(*t*_0_) = *I*_*H*_(*t*_0_) = *D*_*H*_(*t*_0_) = *D*_*C*_(*t*_0_) = *R*_*H*_(*t*_0_) = *R*_*C*_(*t*_0_) = 0.

We estimate the proportion of pre-tested exposed individuals *ρ* is set to be 0.1, and the mortality rate *c* is set to be 0.05 [2, 35]. The basic reproduction rate *R*_0_ is specified based on the government response to the outbreak (the three phases); for instance, if a partial shutdown is implemented at time *t*_1_, and then a full shutdown at *t*_2_, we will have 1 ≤ *R*_0_(*t*_2_ ≤ *t* ≤ 250) ≤ *R*_0_(*t*_1_ ≤ *t* ≤ *t*_2_) ≤ *R*_0_(0 ≤ *t* ≤ *t*_1_). In our case, using the estimates obtained in [21] and depending on the coast being studied, we set the first phase *R*_0_(0 ≤ *t* ≤ *t*_1_) to be either 2.7 or 2.9, the second phase *R*_0_(*t*_1_ ≤ *t* ≤ *t*_2_) to be either 2.3 or 2.5, and the third phase *R*_0_(*t*_2_ ≤ *t* ≤ 250) = 1. The West Coast will take the lower of the two values to account for the lower average population density.

To account for the variations of incubation period *α*^−1^ and infectious period *γ*^−1^ within the population, we sampled 100 values of each of them from an Erlang distribution of shape 2 [21] at each timestep *t*_*i*_. Let us denote these 100 values as 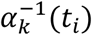 and 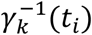, where *k* = 1, …, 100. The Erlang distribution for which we sampled *α*^−1^ from has a mean set to be 5.2 days, and for *γ*^−1^, 3 days, as suggested by [16, 19, 20]. The minimum of the distribution range is also restricted to be 1, i.e. minimum possible incubation period is 1 day. Correspondingly, we will have 100 values of the contact rate *β* for at timestep, *β*_*k*_(*t*_*i*_) = *R*_0_(*t*_*i*_)*γ*_*k*_(*t*_*i*_). We then performed 100 iterations of the fourth-order Runge-Kutta for each of these *α*_*k*_, *β*_*k*_, and *γ*_*k*_ to obtain 100 values for the next timestep, *t*_*i*+1_. Finally, the mean of these 100 values at *t*_*i*+1_ was taken. It is also important to note that at every iteration, the effective total population *N* should be updated.

In summary, our chosen values are organized in the following table.

| Variable | Value (and distribution if applicable) |
| --- | --- |
| $\alpha^{-1}$ | 5.2 days, Erlang distributed with shape 2 |
| $\gamma^{-1}$ | 3 days, Erlang distributed with shape 2 |
| $R_0$ (Phase 1) | 2.7 (West Coast); 2.9 (East Coast) |
| $R_0$ (Phase 2) | 2.3 (West Coast); 2.5 (East Coast) |
| $R_0$ (Phase 3) | 1 |
| $\beta$ | $R_0\gamma$ |
| $\rho$ | 0.1 |
| $c$ | 0.05 |

## Results

Using elements outlined in the previous sections to model the situation for the West Coast, East Coast, as well as the entire US, we ran 50 simulations using the model described in the previous section. We then compared our simulation results to current data between January 20 and March 19 [2, 13], assuming a delay in reporting time of about 8 to 10 days and using population estimates derived from [4].

### The West Coast

The first case of COVID-19 in the West Coast was reported on January 20, 2020 in Seattle, WA. We used a population estimate of 53 million. If January 20 is designated Day 0, we assume Phase 2 started at around Day 45, and Phase 3 started at around Day 60. The simulation results suggest that, under quasi-worst-case-scenario, the data for the number of people infected by COVID-19 is far more than reported: about 80% of *I*_*C*_ (those infected but not isolated) are not accounted for. This is probable as many symptoms of COVID-19 may be passed off as just cases of mild flu. In the quasi-worst-case-scenario, we predict the number of reported infections at its peak to be about 25,000 (Fig. 2) and the actual number of infections at about 90,000 (Fig. 3).

**Figure 2:**
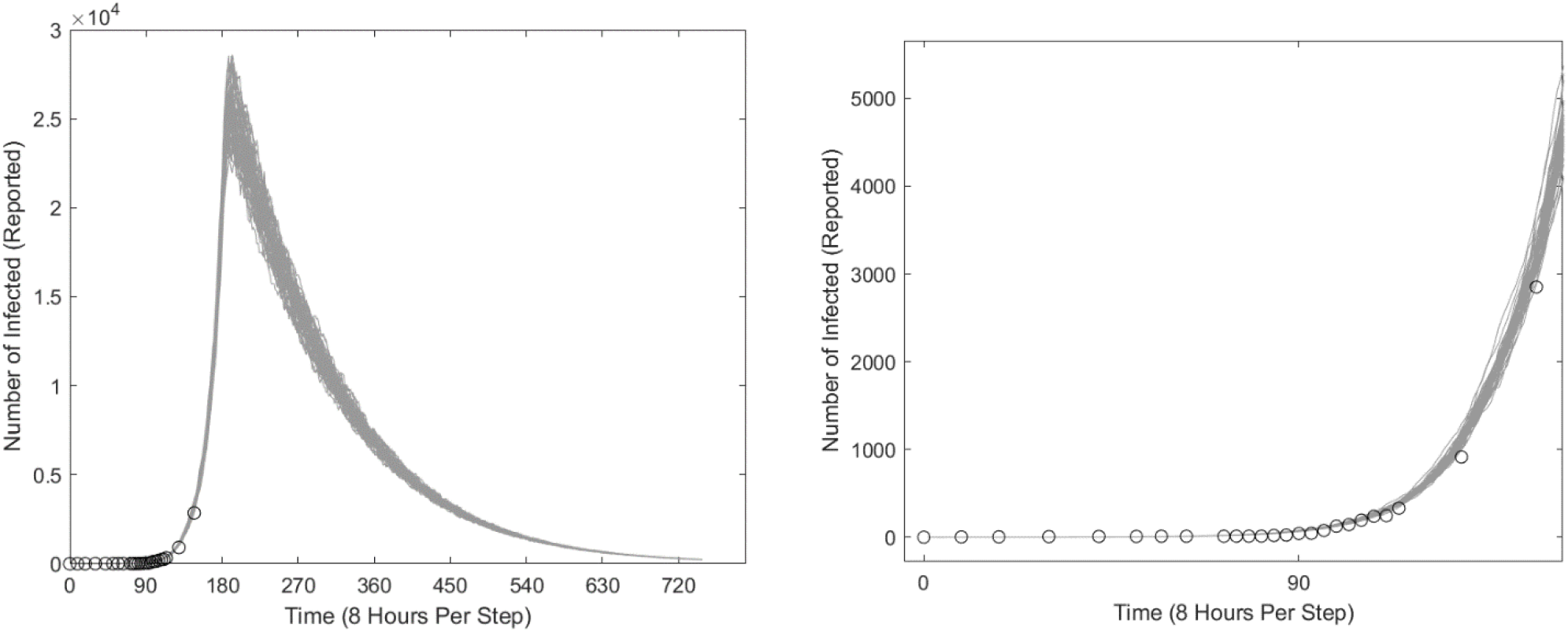
Simulations of COVID-19 outbreak in the West Coast. Each figure comprises 50 different simulations that are represented by the “fuzz” lines. On the x-axis, the starting date at the origin is January 12, 2020, and 90 represents 90 steps of 8 hours. The blue circles represent the actual data of reported cases, adjusted for delay. The left figure estimates the total number of reported infections *I*_*R*_ over time using the formula *I*_*R*_ = *I*_*H*_ + 0.2*I*_*C*_, and the right figure is a magnification that includes available data to date.

**Figure 3:**
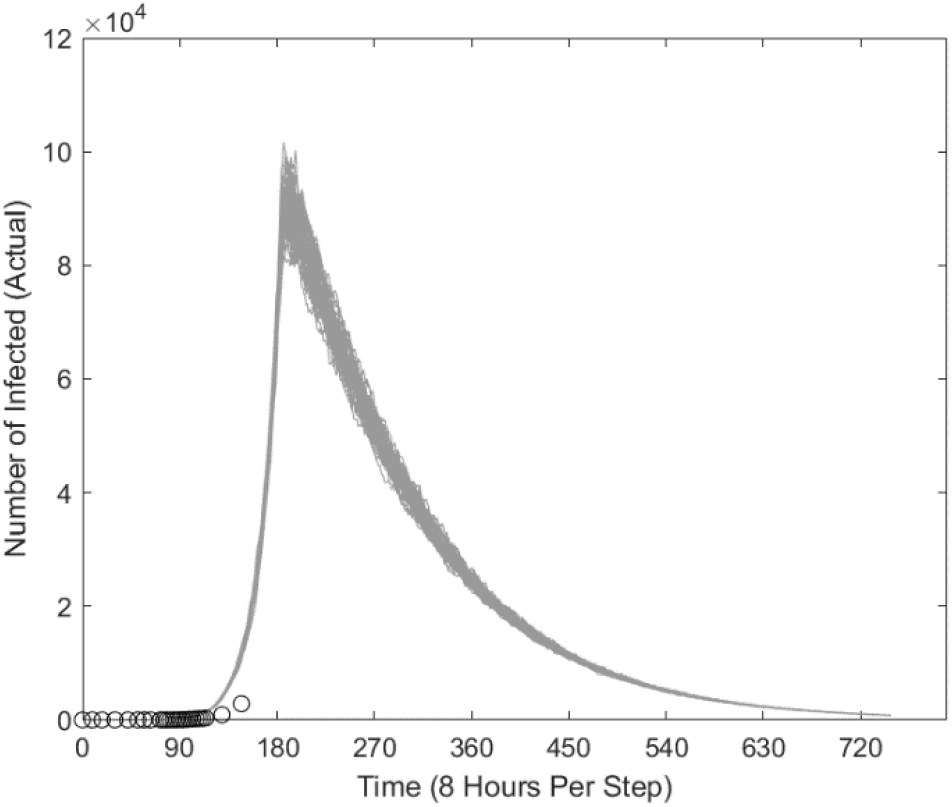
Estimated number of total infections in the West Coast over time, using the formula *I* = *I*_*H*_ + *I*_*C*_. The number of reported cases is much under the actual number.

### The East Coast

The first case of COVID-19 in the East Coast was reported on February 1, 2020 in Boston, MA. However, since no other new cases were reported for about two weeks, we assume a starting time of one month later than for the West Coast. In the graphs below, we will still plot them starting from the first reported case in the US (i.e. January 20). For the East Coast, we used a population estimate of 152 million. Based on government actions, we also assume that the starting dates for Phases 2 and 3 are about a week behind that of the West Coast. The simulation results are similar to that of the West Coast; for a quasi-worst-case scenario, the actual number of infections is predicted to be about 400,000 for reported (Fig. 4), and about 1,450,000 for the actual number of infections (Fig. 5).

**Figure 4:**
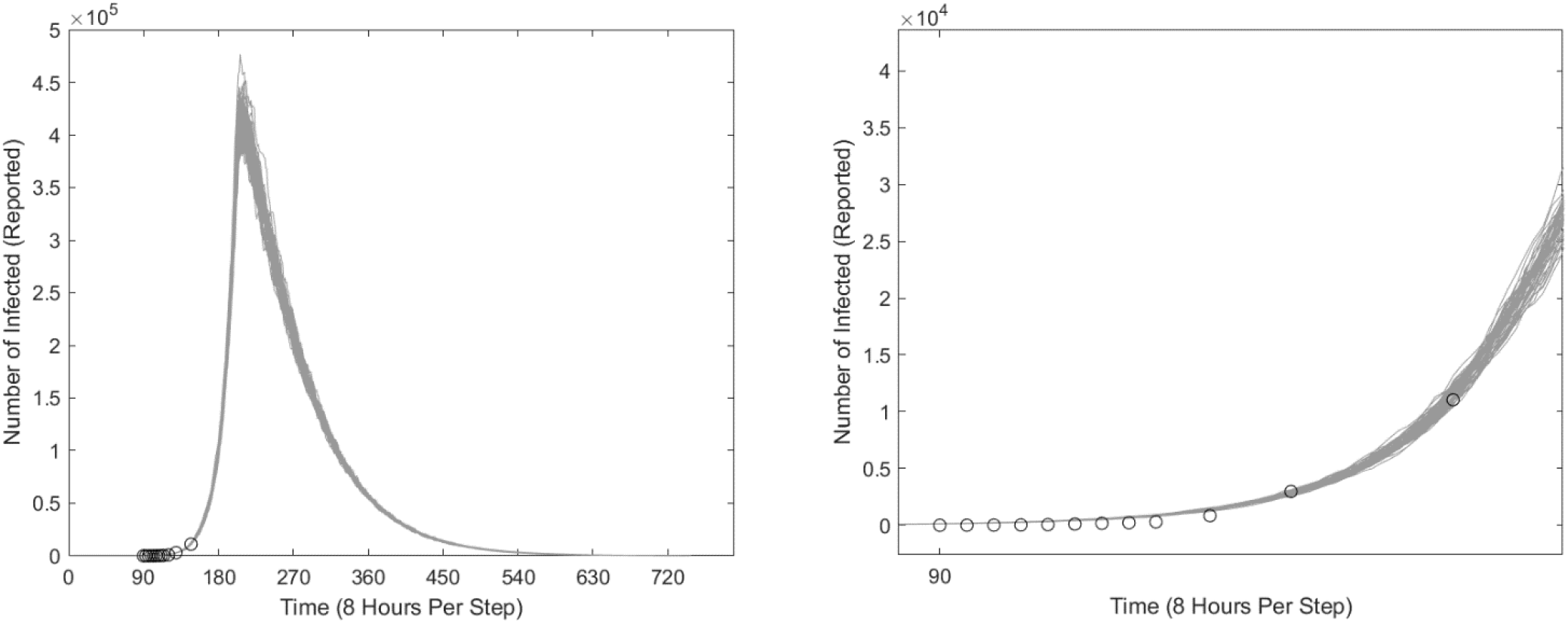
Simulations of COVID-19 outbreak in the East Coast. Each figure comprises 50 different simulations that are represented by the “fuzz” lines. On the x-axis, the starting date at the origin is January 12, 2020, and 90 represents 90 steps of 8 hours. The blue circles represent the actual data of reported cases, adjusted for delay. The left figure estimates the total number of reported infections *I*_*R*_ over time using the formula *I*_*R*_ = *I*_*H*_ + 0.2*I*_*C*_, and the right figure is a magnification that includes available data to date.

**Figure 5:**
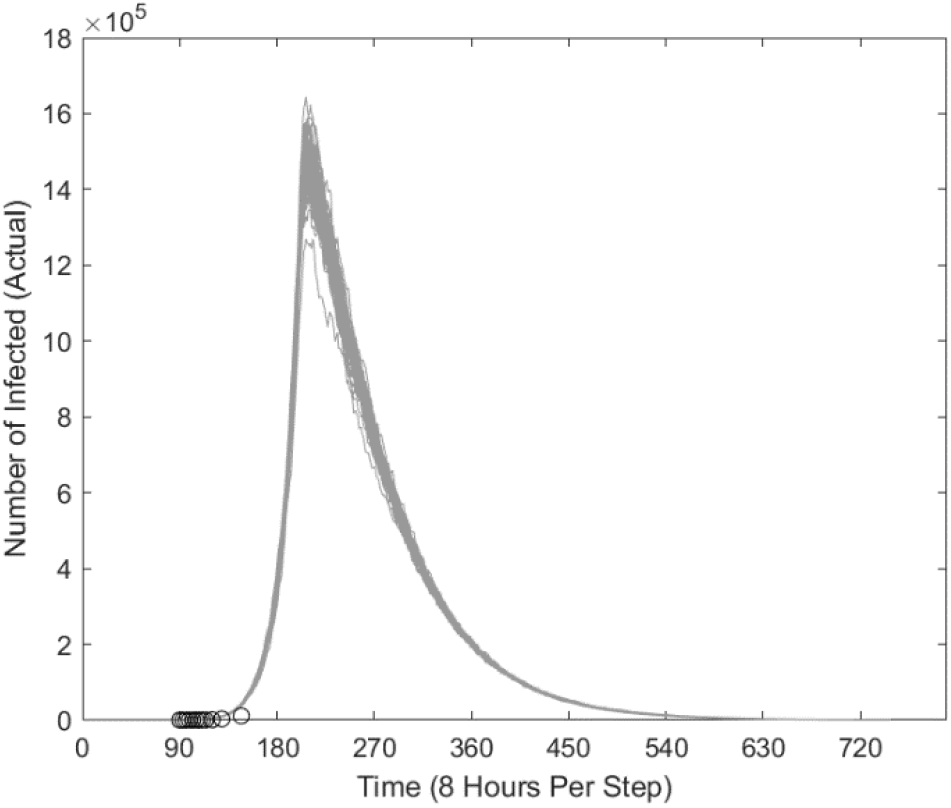
Estimated number of total infections in the East Coast over time, using the formula *I* = *I*_*H*_ + *I*_*C*_. The number of reported cases is much under the actual number.

### The United States: Hypothetical 25% Infected Worse-Case Scenario

As COVID-19 has escalated into a pandemic, we also considered that a greater number of people would fall ill due to the lack of prior immunity. Considering the previous pandemic, the 2009 Influenza A (H1N1) pandemic where around 24% of people were infected [14], as well as the non-static nature of our variables, we ran another simulation on the entire US population with approximately 22% to 26% of the population infected (based on randomness) to predict such a scenario. For this simulation experiment, we used values for and starting days of the phases that was a rough weighted estimate of the two coasts, but used the same values for the other variables as described in the previous section. We also assumed five infected individuals were introduced into the population.

As Fig. 6 illustrates, in this quasi-worst-case-scenario, about 3.8 million of the US population would die from COVID-19 (assuming a death rate of 5%), and the remaining 76.2 million would recover from the virus.

**Figure 6:**
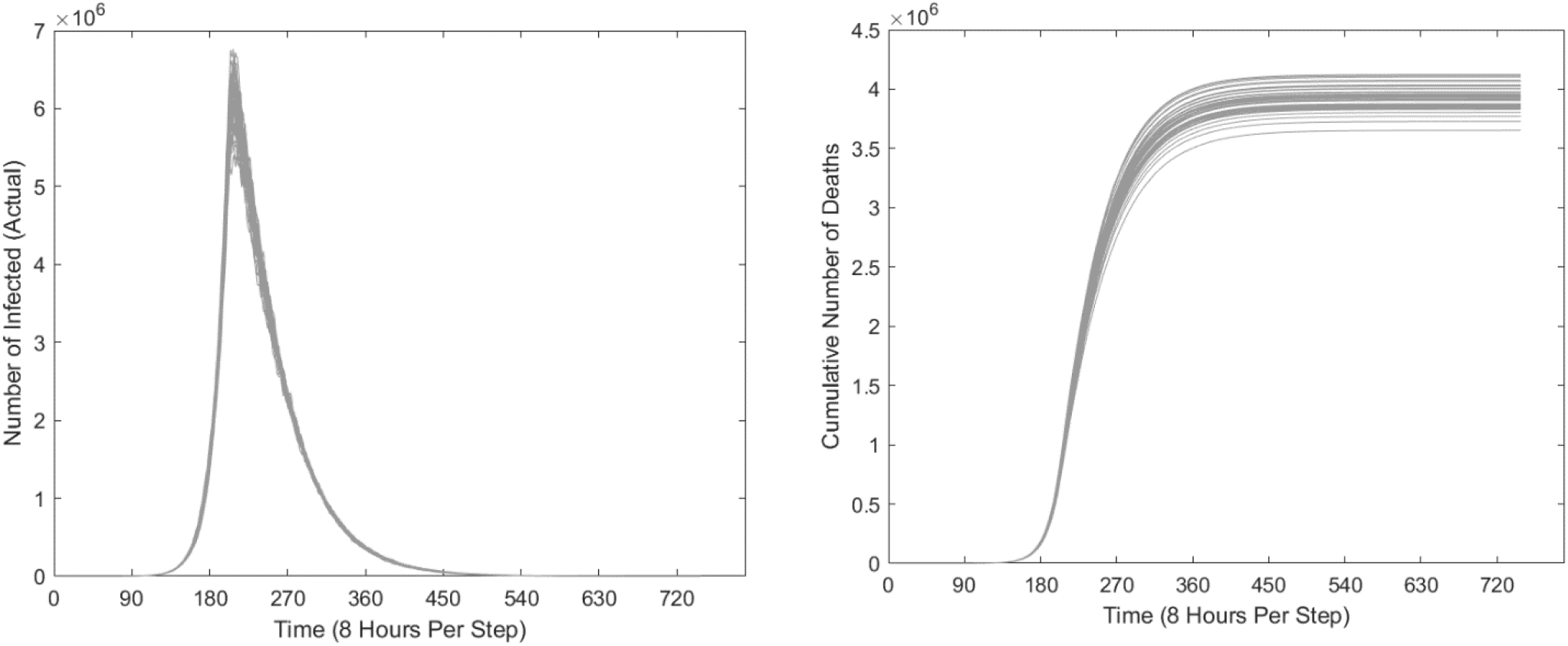
Simulations of a quasi-worst-case COVID-19 outbreak across the US based on a hypothetical scenario where around 25% of the US will be infected. The left figure shows the estimated number of infected people over time, and the right figure estimates the cumulative number of deaths by considering a constant 5% mortality at each unit time.

## Discussion

This paper presents a model for simulating the scale of COVID-19 pandemic in the United States. The simulations focused on the West and East Coasts (Figs. 2-5), and also modeled a hypothetical worst-case scenario for the entire US (Fig. 6). The model implemented factors specific to the US, which include the three phases of government response, travel restrictions that restrict cross-boundary transmissions, and limits to testing kits and healthcare accessibility resulting in semi-strict isolation.

The model fits the reported data by assuming that about 80% of the non-isolated cases are not reported (Figs. 2, 4); however, the actual number of cases may be much higher (Figs. 3, 5). That assumption agrees with a prior study [17] and provides another evidence that COVID-19 is not easy to contain. The model also predicts that the peak of the outbreak would occur by early April, and the outbreak would wind down by the start of July. However, the model does not account for delays in reporting; as such, one should expect the peak reported number of infections to occur around mid-April. In addition, the peak would occur sooner on the West Coast, which was affected first. However, all peaks should occur around mid-April unless there are major changes to the reproduction rates, for which there is no current evidence. It should be noted that the trends in the model’s estimates share similarities with the epidemic curves during the SARS and MERS outbreaks [32, 34].

One way to mitigate the spread of COVID-19 is to successfully isolate more infected people, i.e. to have a higher relative proportion of *I*_*H*_ over *I*_*C*_. That outcome should be achieved when defective testing kits are replaced and more testing kits are allocated, so that more exposed people can be tested and isolated accordingly. For instance, if 50% more people had been tested (*ρ* = 0.15) since the start of the outbreak, the model would predict 40% of the projected severity (Figs. 7, 8). As the US faced mishaps in the allocation of testing kits during the initial handling of COVID-19, the original peak is likely inevitable. Testing more people from this point onwards (i.e. late March) would accelerate the cool down; a simulation for *ρ* = 0.3 is shown in Figs. 9, 10.

**Figure 7:**
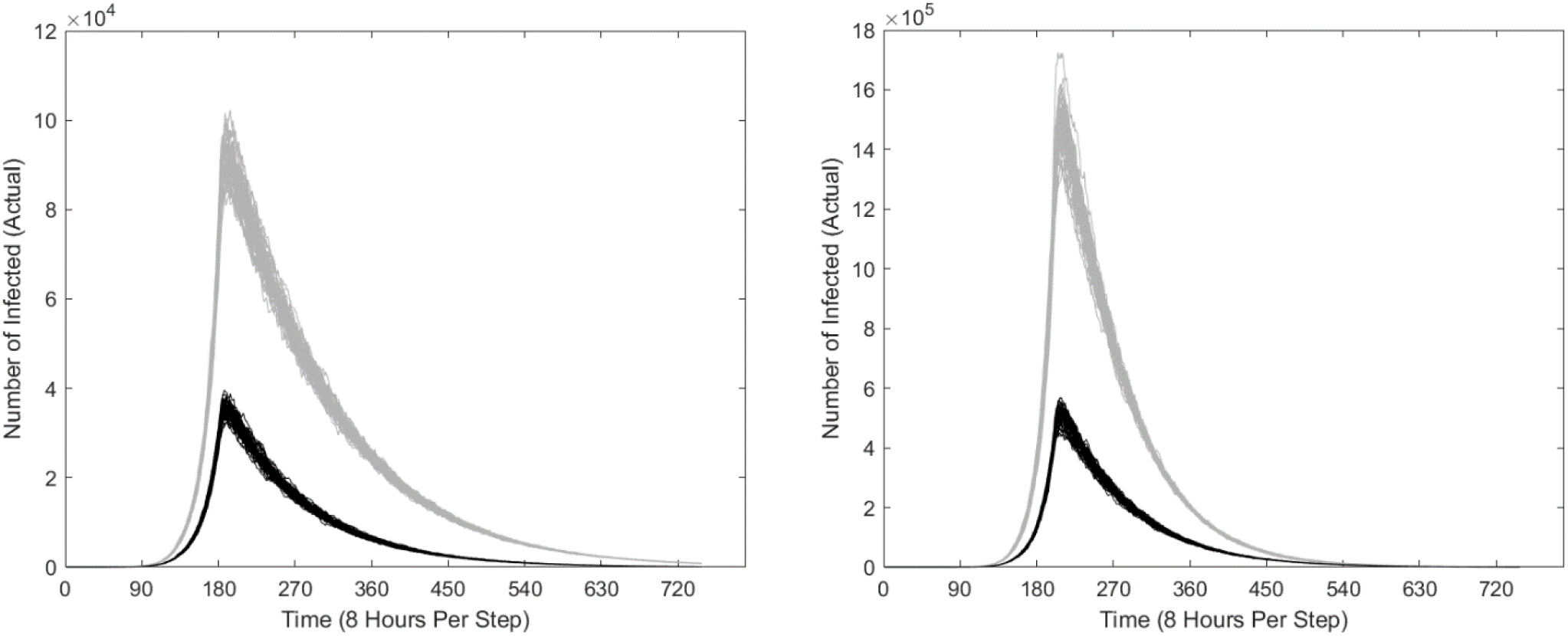
Comparison of COVID-19 outbreak with 50% more people tested since the onset (solid) versus the current situation (transparent) by considering the total number of infected. (*I* = *I*_*H*_ + *I*_*C*_). The left and right figures show the West and East Coasts, respectively.

**Figure 8:**
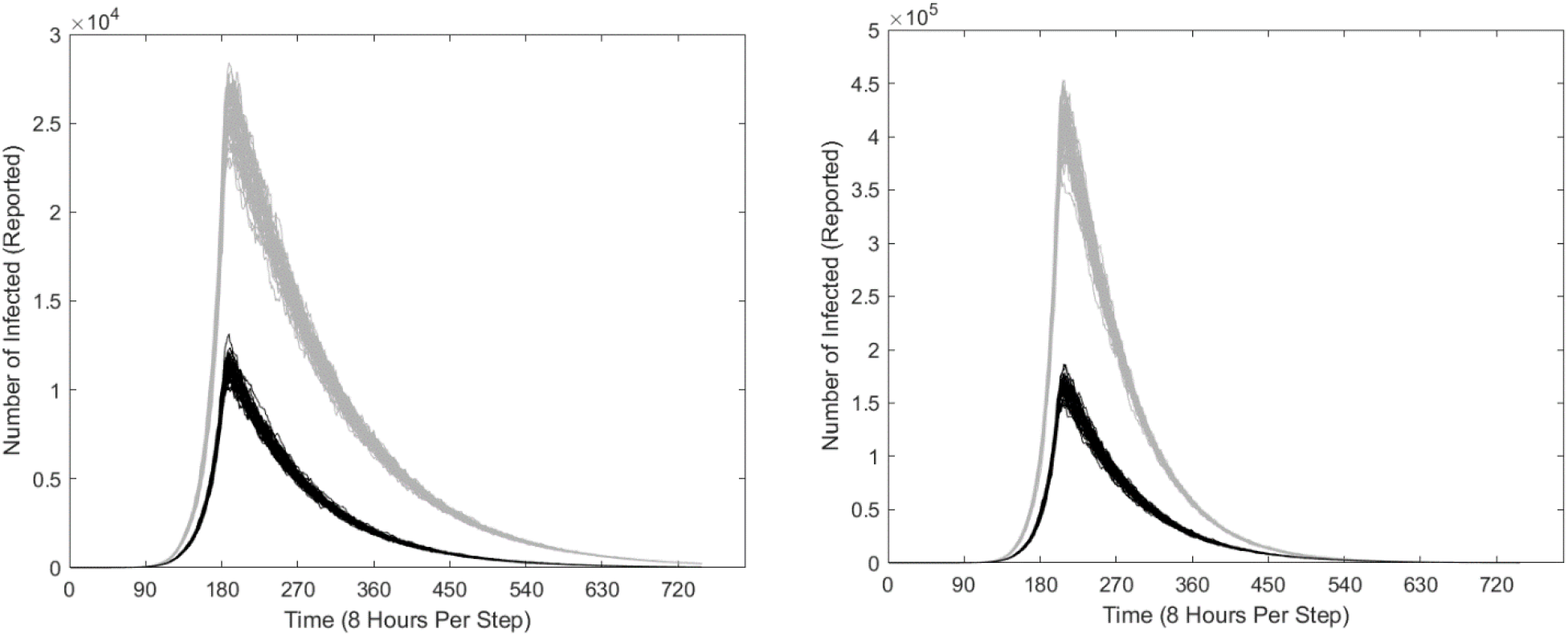
Comparison of COVID-19 outbreak with 50% more people tested since the onset (solid) versus the current situation (transparent) by considering only the infected that are reported. (*I*_*R*_ = *I*_*H*_ + 0.2*I*_*C*_). The left and right figures show the West and East Coasts, respectively.

**Figure 9:**
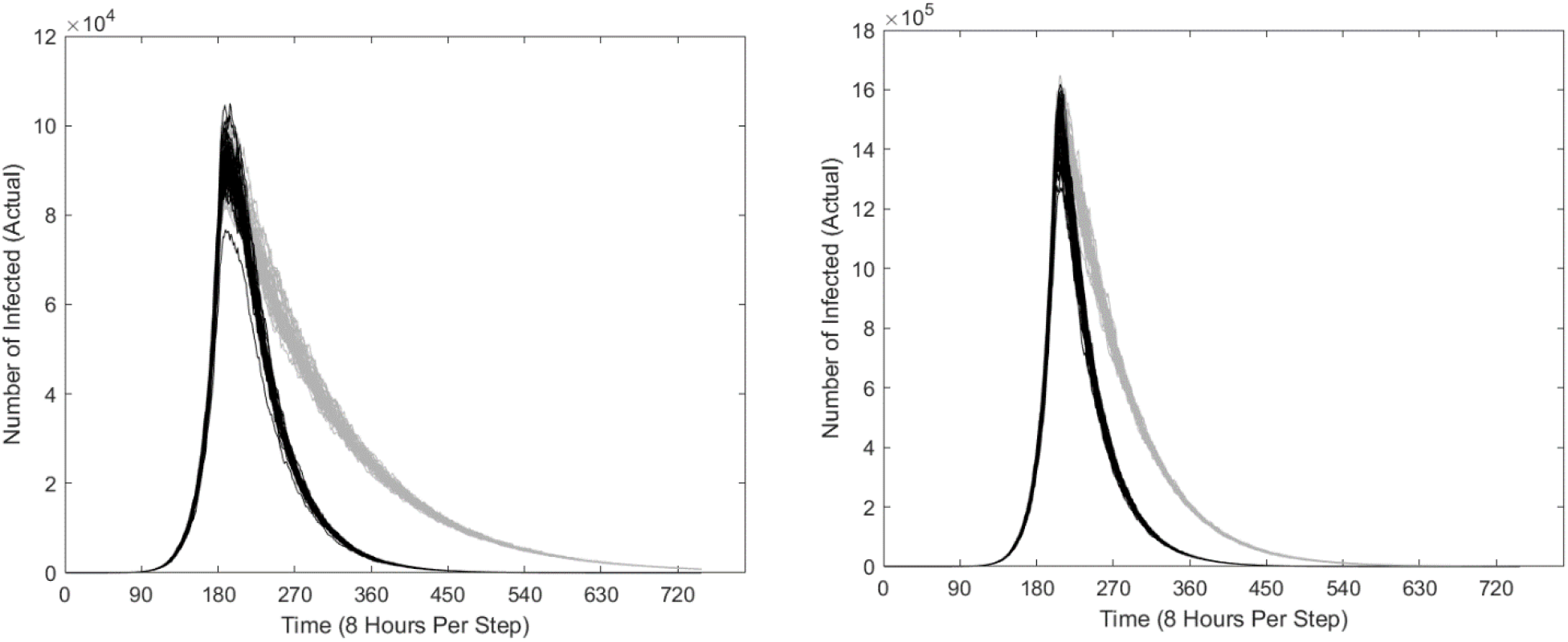
Comparison of COVID-19 outbreak with 3X number of people tested starting late March (solid) versus the current situation (transparent) by considering the total number of infected. (*I* = *I*_*H*_ + *I*_*C*_). The left and right figures show the West and East Coasts, respectively.

**Figure 10:**
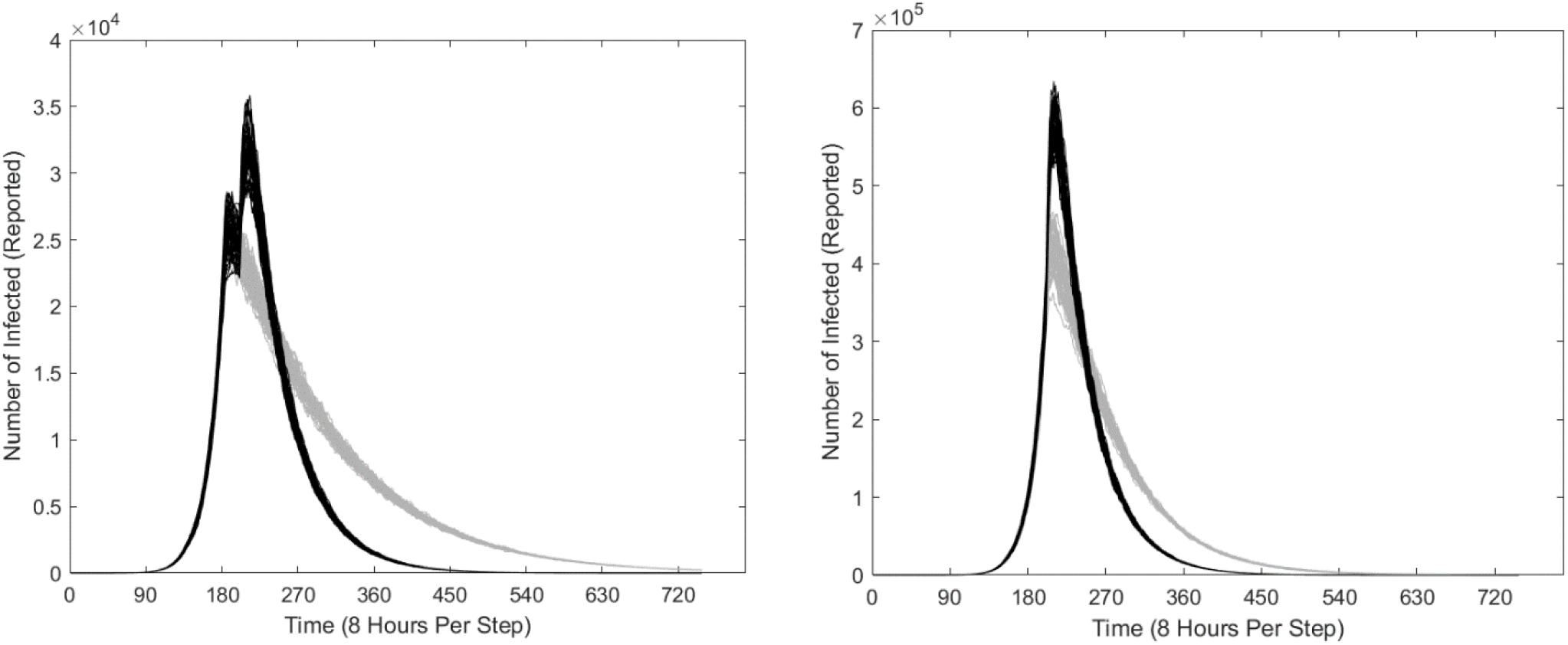
Comparison of COVID-19 outbreak with 3X number of people tested starting late March (solid) versus the current situation (transparent) by considering only the infected that are reported. (*I*_*R*_ = *I*_*H*_ + 0.2*I*_*C*_). The left and right figures show the West and East Coasts, respectively. Note that more reported cases are initially observed than originally projected because testing many more people shifts potentially unreported *I*_*C*_ to the *I*_*H*_ cohort. Two peaks are observed for the West Coast: the original peak is projected have already passed, and implementing 3X more tests will result in another peak of reported infections.

Additional ways to mitigate the mortality of COVID-19 include improving accessibility to healthcare and boosting quantities of essential supplies such as respirators and ventilators. However, the sudden onset of COVID-19 and the overall unpreparedness of the United States make these data inaccessible. It should also be noted that while the global mortality during COVID-19 is currently 3.8%, it is noticeably different across nations [24]. As a result, we chose a 5% mortality rate (*c* = 0.05) in our quasi-worst-case scenario prediction of COVID-19 in the US.

Other factors at a molecular level can further modulate our variables. For instance, the actual *R*_0_ in the US might be different as the transmissibility and infectivity of CoVs have been shown to be modulated by abiotic factors such as temperature and humidity [1], and the robustness of host immunity further affects the etiology, pathogenesis, and overall virulence of CoVs [9]. As a result, our paper used numbers from prior studies that are based on other countries hit relatively hard by COVID-19 such as China, South Korea, and Italy, and enables us to successfully construct a quasi-worst-case model of COVID-19 in the US.

We are mindful that computational simulations are, by their very nature, approximations. There are currently no predictive models that satisfactorily produce a picture of the spread or clinical impact of the disease as too many variables can affect the spread of a disease, especially for pandemics like COVID-19. Moreover, simulations can vary noticeably with small variations in assumptions and parameters. To cite one example, minor changes in the distribution of the infected population *I* between *I*_*H*_ and *I*_*C*_ can dramatically affect the model’s predictions. Also, at the societal level, the length of time that the cohort of patients remain hospitalized is often unknown in the early stages of a pandemic, and can greatly influence the use and deployment of medical supplies and personnel, thus altering the course of the infection and recovery rates. Nevertheless, the few scenarios presented and discussed in this paper strongly suggest that even with current containment and mitigation efforts, the COVID-19 outbreak will significantly impact the health of the US population.

## Data Availability

All data used are available publicly from Kaggle and CDC.

## Acknowledgements

We thank Professor Bruce Ganem (Department of Chemistry and Chemical Biology) and Professor John Muckstadt (School of Operations Research and Industrial Engineering) of Cornell University for their encouragement, invaluable advice and dedicated guidance throughout the study.

## Author Contributions

YYY conceived the study and obtained the data. YRY and WJY designed and implemented the model and interpreted the results while YYY provided biological context. YYY, YRY and WJY wrote the manuscript. Authors contributed equally and should be considered as co-first authors.

## Conflict of Interests

The authors declare that they have no competing interests.

## Notes

### Competing Interest Statement

The authors have declared no competing interest.

### Funding Statement

No funding was received for the study.

## References

1. Casanova LM, Jeon S, Rutala WA, Weber DJ, Sobsey MD. Effects of Air Temperature and Relative Humidity on Coronavirus Survival on Surfaces. 2010. Applied and Environmental Microbiology 76:2712–2717.

2. Centers for Disease Control and Prevention. 2020. Coronavirus Disease 2019 (COVID-19) Situation Summary. Available from https://www.cdc.gov/coronavirus/2019-ncov/cases-updates/summary.html

3. Centers for Disease Control and Prevention. 2020. International Locations with Confirmed COVID-19 Cases. Available from https://www.cdc.gov/coronavirus/2019-ncov/cases-updates/world-map.html

4. The United States Census Bureau. 2020, City and Town Population Totals: 2010-2018. Available from https://www.census.gov/data/tables/time-series/demo/popest/2010s-total-cities-and-towns.html

5. Cui J, Li F, Shi ZL. Origin and evolution of pathogenic coronaviruses. 2019. Nature Reviews Microbiology 17:181–192.

6. Drosten C, Günther S, Preiser W, van der Werf S, Brodt HR, Becker S, et al. Identification of a Novel Coronavirus in Patients with Severe Acute Respiratory Syndrome. 2003. New England Journal of Medicine 348:1967–1976.

7. Dureau J, Kalogeropoulos K, Baguelin M. Capturing the time-varying drivers of an epidemic using stochastic dynamical systems. 2013. Biostatistics 14:541–555.

8. Fehr AR, Perlman, S. Coronaviruses: An Overview of Their Replication and Pathogenesis. In: Maier HJ, Bickerton E, Britton P, editors. Coronaviruses: Methods and Protocols. Springer, New York, NY; 2015. pp. 1–23

9. Frieman M, Heise M, Baric, R. SARS coronavirus and innate immunity. 2008. Virus Research 133(1):101–112.

10. Guan W, Ni ZY, Hu Y, Liang WH, Ou CQ, He JX, et al. et al. Clinical Characteristics of Coronavirus Disease 2019 in China. 2019. New England Journal of Medicine doi: 10.1056/NEJMoa2002032.

11. Holshue ML, DeBolt C, Lindquist S, Lofy KH, Weisman J, Bruce H, et al. First Case of 2019 Novel Coronavirus in the United States. 2020. New England Journal of Medicine 382:929–936.

12. Hubbard J, West B. Differential Equations: A Dynamical Systems Approach. Springer, New York, NY; 1997.

13. Kaggle. Novel Corona Virus 2019 Dataset. 2020. Available from https://www.kaggle.com/sudalairajkumar/novel-corona-virus-2019-dataset

14. Kerkhove MDV, Hirve S, Koukounari A, Mounts AW. Estimating age-specific cumulative incidence for the 2009 influenza pandemic: a meta-analysis of A(H1N1)pdm09 serological studies from 19 countries. 2013. Influenza and Other Respiratory Viruses 7:872–886.

15. Ksiazek TG, Erdman D, Goldsmith CS, Zaki SR, Peret T, Emery S, et al. A Novel Coronavirus Associated with Severe Acute Respiratory Syndrome. 2003. New England Journal of Medicine 348:1953–1966.

16. Lauer SA, Grantz KH, Bi Q, Jones FK, Zheng Q, Meredit HR, et al. The Incubation Period of Coronavirus Disease 2019 (COVID-19) From Publicly Reported Confirmed Cases: Estimation and Application. 2020. Annals of Internal Medicine doi: 10.7326/M20-0504

17. Li R, Pei S, Chen B, Song Y, Zheng T, Yang W, et al. Substantial undocumented infection facilitates the rapid dissemination of novel coronavirus (SARS-CoV2). 2020. Science doi: 10.1126/science.abb3221

18. Lin Q, Zhao S, Gao D, Lou Y, Yang S, Musa SS, et al. A conceptual model for the coronavirus disease 2019(COVID-19) outbreak in Wuhan, China with individual reaction and governmental action. 2020. International Journal of Infectious Diseases 93:211–216.

19. Linton NM, Kobayashi T, Yang Y, Hayashi K, Akhmetzhanov AR, Jung SM, et al. Incubation Period and Other Epidemiological Characteristics of 2019 Novel Coronavirus Infections with Right Truncation: A Statistical Analysis of Publicly Available Case Data. 2020. Journal of Clinical Medicine 9(2):538.

20. Liu T, Hu J, Xiao J, He G, Kang M, Rong Z, et al. Time-varying transmission dynamics of Novel Coronavirus Pneumonia in China. 2020. bioRxiv doi: 10.1101/2020.01.25.919787.

21. Liu T, Hu J, Xiao J, He G, Kang M, Rong Z, et al. Transmission dynamics of 2019 novel coronavirus. 2020. bioRxiv doi: 10.1101/2020.01.25.919787v1.full

22. Liu Y, Gayle AA, Wilder-Smith A, Rocklöv J. The reproductive number of COVID-19 is higher compared to SARS coronavirus. 2020. Journal of Travel Medicine 27:taaa021.

23. Lu R, Zhao X, Li J, Niu P, Yang B, Wu H, et al. Genomic characterisation and epidemiology of 2019 novel coronavirus: implications for virus origins and receptor binding. 2020. The Lancet 395:565–574.

24. Roser M, Ritchie H, Ortiz-Ospina E. Coronavirus Disease (COVID-19) – Statistics and Research. 2020. Available from https://ourworldindata.org/coronavirus

25. Science Magazine. Wuhan seafood market may not be source of novel virus spreading globally. 2020. Available from https://www.sciencemag.org/news/2020/01/wuhan-seafood-market-may-not-be-source-novel-virus-spreading-globally

26. Su S, Wong G, Shi W, Liu J, Lai ACK, Zhou J, et al. Epidemiology, Genetic Recombination, and Pathogenesis of Coronaviruses. 2016. Trends in Microbiology 24:490–502.

27. The New York Times. Coronavirus Test Kits Sent to States Are Flawed, C.D.C. Says. 2020. Available from https://www.nytimes.com/2020/02/12/health/coronavirus-test-kits-cdc.html

28. The New York Times. With Test Kits in Short Supply, Health Officials Sound Alarms. 2020. Available from https://www.nytimes.com/2020/03/06/health/testing-coronavirus.html

29. The New York Times. Estimates Fall Short of F.D.A.’s Pledge for 1 Million Coronavirus Tests. 2020. Available from https://www.nytimes.com/2020/03/03/health/coronavirus-tests-fda.html

30. The New York Times. There Aren’t Enough Ventilators to Cope With the Coronavirus. 2020. Available from https://www.nytimes.com/2020/03/18/business/coronavirus-ventilator-shortage.html

31. The New York Times. US Virus Testing Faces New Headwind: Lab Supply Shortages. 2020. Available from https://www.nytimes.com/aponline/2020/03/20/health/ap-us-med-virus-outbreak-us-testing.html

32. Wallinga J, Teunis P. Different Epidemic Curves for Severe Acute Respiratory Syndrome Reveal Similar Impacts of Control Measures. 2004. American Journal of Epidemiology 160:509–516.

33. Wang M, Qi J. A deterministic epidemic model for the emergence of COVID-19 in China. 2020. medRxiv doi:10.1101/2020.03.08.20032854.

34. World Health Organization. MERS-CoV maps and epicurves. 2020. Available from http://www.who.int/emergencies/mers-cov/maps-september-2017/en/

35. World Health Organization. Novel Coronavirus (2019-nCoV) situation reports. 2020. Available from https://www.who.int/emergencies/diseases/novel-coronavirus-2019/situation-reports

36. Zaki AM, van Boheemen S, Bestebroer TM, Osterhaus ADME, Fouchier RAM. Isolation of a Novel Coronavirus from a Man with Pneumonia in Saudi Arabia. 2012. New England Journal of Medicine 367:1814–1820.

37. Zhu N, Zhang D, Wang W, Li X, Yang B, Song J, et al. A Novel Coronavirus from Patients with Pneumonia in China. 2019. New England Journal of Medicine 382:727–733.

38. Zhuang Z, Zhao S, Lin Q, Cao P, Lou Y, Yang L, et al. Preliminary estimating the reproduction number of the coronavirus disease (COVID-19) outbreak in Republic of Korea and Italy by 5 March 2020. 2020. medRxiv doi:10.1101/2020.03.02.20030312.

